# Large Language Models (LLMs) and Empathy – A Systematic Review

**DOI:** 10.1101/2023.08.07.23293769

**Authors:** Vera Sorin, Danna Brin, Yiftach Barash, Eli Konen, Alexander Charney, Girish Nadkarni, Eyal Klang

## Abstract

**Purpose:** Empathy, a cornerstone of human interaction, is a unique quality to humans that Large Language Models (LLMs) are believed to lack. Our study aims to review the literature on the capacity of LLMs in demonstrating empathy

**Methods:** We conducted a literature search on MEDLINE up to July 2023. Seven publications ultimately met the inclusion criteria.

**Results:** All studies included in this review were published in 2023. All studies but one focused on ChatGPT-3.5 by OpenAI. Only one study evaluated empathy based on objective metrics, and all others used subjective human assessment. The studies reported LLMs to exhibits elements of empathy, including emotions recognition and providing emotionally supportive responses in diverse contexts, most of which were related to healthcare. In some cases, LLMs were observed to outperform humans in empathy-related tasks.

**Conclusion:** LLMs demonstrated some aspects of empathy in variable scenarios, mainly related to healthcare. The empathy may be considered “cognitive” empathy. Social skills are a fundamental aspect of intelligence, thus further research is imperative to enhance these skills in AI.

## Introduction

Empathy, a fundamental aspect of human interaction, can be characterized as the ability to experience the emotions of another being within oneself. The origin of the word “empathy” dates back to the 1880s, when Theodore Lipps determined the word “einfuhlung” (“in-feeling”) to describe the emotional appreciation of another’s feelings (1). Empathy involves recognition of others’ feelings, the causes of these feelings, and the ability to participate in an emotional experience of an individual without becoming part of it (1). In the context of healthcare, empathy enables health care professionals and patients to communicate. It is described as “the ability to see the world through someone else’s eyes”, having the ability to imagine what someone else is thinking and feeling in a given situation (2).

Large Language Models (LLMs) have demonstrated remarkable capabilities across various tasks, including text summarization, question-answering, and text generation (3). There are numerous studies on potential applications in healthcare, as an educational tool and as a support tool in clinical work (4, 5). Some publications suggest that despite impressive natural language processing abilities, LLMs lack empathy, a quality that is unique to humans (6-9).

Few studies in the literature, discuss and evaluate LLMs performance in tasks associated with emotional intelligence, theory of mind, and empathy. Thus, the aim of our study was to systematically review the literature on the capacity of LLMs in demonstrating empathy.

## Methods

We searched the literature on LLMs and empathy using MEDLINE. Studies published up to July 2023 were included. The search query was “((“large language models”) OR (llms) OR (gpt) OR (chatgpt)) AND ((empathy) OR (“emotional awareness”) OR (“emotional intelligence”) OR (emotion))”. The initial search yielded 34 studies. We also searched the references lists of relevant studies, including some key studies from major medical journals, for any additional studies that may have been missed during the initial search. This process resulted in the retrieval of additional three studies.

The inclusion criteria for our study were English language full-length publications that evaluated empathy in LLMs outputs. Excluded were papers evaluating other topics related to emotional intelligence that were not specifically empathy. For example, papers focusing on “theory of mind” were excluded if they did not specifically address empathy. Two reviewers (VS, EK) independently performed the search and screened the titles and abstract of the articles resulting from the search. Differences in search results were resolved through discussion to reach a consensus. The reviewers then screened selected articles’ full-text for final inclusion. Ultimately, a total of seven publications were included in this review. **Figure 1** presents a flow diagram of the screening and inclusion process.

**Figure 1.**
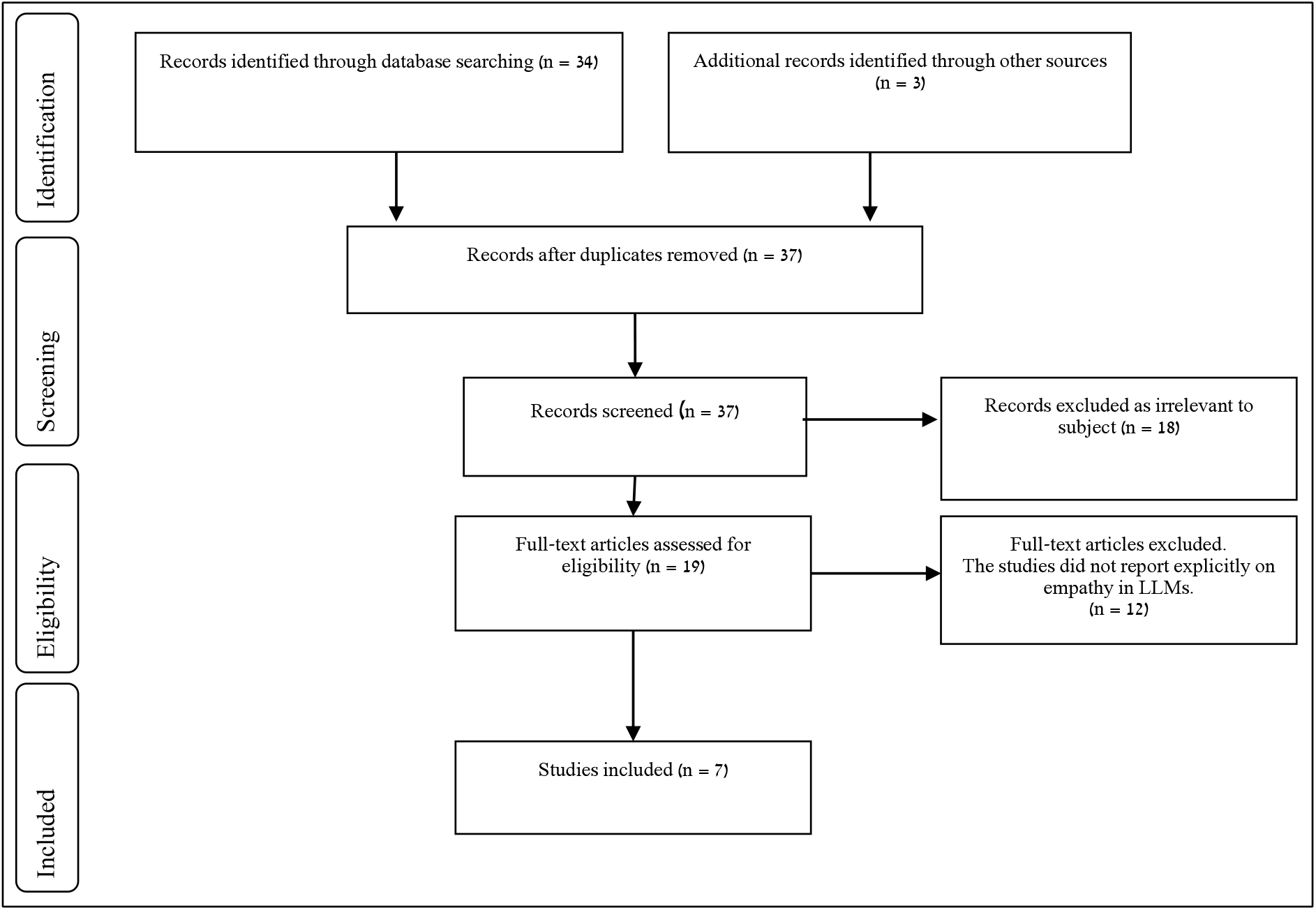
Flow Diagram of the Inclusion Process. Flow diagram of the search and inclusion process based on the Preferred Reporting Items for Systematic Reviews and Meta-Analyses (PRISMA) guidelines

## Results

All seven studies included in this review were published in 2023. Despite the emergence of various LLMs, all studies but one focused on ChatGPT-3.5 by OpenAI. Six out of seven studies evaluated ChatGPT’s empathy based on subjective human evaluation, as opposed to objective metrics. All studies but one evaluated empathy in ChatGPT in medical context. The results of the studies included are summarized in **Table 1**.

**Table 1.** Studies Evaluating Aspects of Empathy Exhibited by Large Language Models.

| Study (ref) | Year | Journal | Objective | LLM Model | Key Findings | Limitations of LLMs |
| --- | --- | --- | --- | --- | --- | --- |
| Webb et al. (10) | 2023 | Cureus | Breaking bad news in emergency medicine | ChatGPT-3.5 | ChatGPT facilitated realistic scenario design, active roleplay, and effective feedback through the application of the SPIKES framework for breaking bad news. | Specific limitations were not discussed. General limitations discussed include limited training dataset, performance being influenced by prompts design, potential for inaccurate responses and inability to convey parts of human communications such as eye contact, pausing to listen, and tone. |
| Ayers et al. (12) | 2023 | JAMA Intern Med | Empathetic responses to patient questions | ChatGPT-3.5 | ChatGPT responses were preferred by evaluators over physicians in 78.6% evaluations and were rated of significantly higher quality and empathy. | Not specifically discussed. ChatGPT tended to provide more lengthy responses, which could potentially be erroneously associated with greater empathy. The study did not assess the chatbot responses for accuracy or fabricated information. |
| Chen et al.<br>(14) | 2023 | arXiv | Simulating psychiatrists and patients in clinical psychiatric scenarios, and evaluating the expression of empathy in the interactions | Chatbots based on ChatGPT | ChatGPT-powered chatbots showed feasibility in simulating some aspects of empathy in psychiatric interactions | The chatbots sometimes forgot initial instructions and showed excessive repetition of general empathy phrases. Additionally, they asked fewer in-depth questions about the symptoms compared to human doctors, potentially affecting their ability to fully understand the patient's condition. The patient chatbots reported symptoms inaccurately. |
| Zhao et al.<br>(17) | 2023 | arXiv | Evaluate emotional dialogue understanding and generation and compare to other supervised models. | ChatGPT-3.5 | Supervised models surpassed ChatGPT in emotion recognition. ChatGPT produced longer responses, but responses were also more specific to the context of the conversation compared to other models. | ChatGPT generated longer responses, and deviated from reference responses when evaluated based on word overlap metrics. It also demonstrated limited understanding of some labels. ChatGPT did not adhere to the same guidelines to determine emotions as were used for the annotated data for the supervised models. |
| Yeo et al. (11) | 2023 | Clin Mol Hepatol | Emotional support for cirrhosis and HCC patients | ChatGPT-3.5 | ChatGPT emulated empathetic responses and offered actionable recommendations for patients and caregivers. | In total, ChatGPT provided comprehensive answers in less than 50% of questions. |
| Elyoseph et al. (16) | 2023 | Front. Psychol | Emotional awareness performance compared to the general population norms | ChatGPT-3.5 | ChatGPT demonstrated significantly higher emotional awareness performance than general population norms, with potential improvements over time | Not specifically discussed. |
| Liu et al. (13) | 2023 | medRxiv | Fine-tuning an LLM to generate responses to patient questions | LLM based on LLaMA-65B; ChatGPT-3.5, GPT-4 | GPT-4 and ChatGPT-3.5 outperformed their model. Interestingly, all language models outperformed physician-generated responses significantly. | One of the reviewer indicated “excessive empathy” in the fine-tuned LLM model. Responses generated by GPT models were considered too lengthy and required a relatively high reading level. |

Ref= reference; LLM= large language model

Empathy is essential in medicine, particularly when breaking bad news to patients. It allows physicians to deliver difficult information in a manner that respects the patient’s emotions and perspective. Webb (10) used ChatGPT to simulate a role-play of breaking bad news in the emergency department. The chatbot successfully set up a training scenario, role-played as a patient, and provided clear feedback through the application of the SPIKES (Setting up, Perception, Invitation, Knowledge, Emotions with Empathy, and Strategy or Summary) framework for breaking bad news (10). In another study, Yeo et al. (11) tested ChatGPT’s ability to provide emotional support to patients diagnosed with hepatocellular carcinoma, and their caregivers. ChatGPT was able to acknowledge the likely emotional response of the patient to their diagnosis. Furthermore, the chatbot provided clear and actionable starting points for a newly diagnosed patient, and offered motivational responses encouraging proactive steps. For caregivers, ChatGPT provided psychological and practical recommendations (11).

Ayers et al. (12) compared the quality and empathy of responses given by ChatGPT and physicians to 195 randomly drawn patient questions from a social media forum. The study found that patients preferred the chatbot’s responses over physician responses in 78.6% of cases. ChatGPT’s responses were rated significantly higher for both quality and empathy, while physician responses were 41% less empathetic than the chatbot responses. The authors noted that ChatGPT tended to provide more lengthy responses, which could potentially be erroneously associated with greater empathy. They concluded that the chatbot may have potential in aiding drafting responses to patient questions (12).

Another study also assessed empathy in chatbot’s responses to patient’s questions. Liu et al. (13) developed a model based on a pre-trained LLaMA-65B and fine-tuned to generate physician-like responses that are professional and empathetic. They evaluated the model on ten actual patient questions in primary care, and compared the responses to those generated by ChatGPT-3.5 and GPT-4, rating them based on empathy, responsiveness, accuracy and usefulness. When evaluating empathy, GPT-4 and ChatGPT-3.5 outperformed their model. Interestingly, all language models outperformed physician-generated responses significantly (13).

Understanding and addressing patients’ emotions is fundamental in mental health. Chen et al. (14) used ChatGPT-powered chatbots to simulate psychiatrists and patients in clinical psychiatric scenarios. The chatbots showed potential in simulating some aspects of empathy. However, they sometimes forgot initial instructions and repeated general empathy phrases too often. They also asked fewer in-depth questions about symptoms compared to physicians, potentially affecting their ability to fully understand the patient’s condition. When simulating patients, the chatbots reported symptoms inaccurately (14).

The Levels of Emotional Awareness Scale (LEAS) is a psychological tool that assesses an individual’s capacity to identify and describe emotions in themselves and others, a fundamental aspect of empathy (15). Elyoseph et al. (16) compared the LEAS score of ChatGPT to the general population norms. They found that ChatGPT demonstrated significantly higher emotional awareness performance. When repeating the test following one month interval, the chatbot’s performance further improved, almost reaching the maximum possible LEAS score. The authors propose that ChatGPT could be helpful for cognitive training of people with emotional awareness impairment, as well as for psychiatric assessment support (16).

Zhao et al. (17) compared ChatGPT to supervised models in terms of emotional dialogue understanding and generation. The tasks they assessed included emotion recognition, emotion cause recognition, dialog act classification, empathetic response generation, and emotional support conversation. The authors found that while supervised models surpassed ChatGPT in emotion recognition, ChatGPT produced longer, more diverse, and context-specific responses, especially when interacting with users in negative emotional states. Interestingly, Zhao et al. (17) also observed a repetitive pattern in ChatGPT’s empathy expressions, similar to the results described by Chen et al (14).

## Discussion

This review shows that LLMs exhibit aspects of empathy, including recognition of emotions, and generation of emotionally supportive responses. Most studies evaluated empathy exhibited in LLMs in variable medical tasks. Some of the studies pointed out potential pitfalls in assessing empathy of LLMs, such as mistaking lengthy responses for increased empathy. Other challenges discussed involve the absence of human communication aspects in LLMs such as eye contact and tone of voice. Limitations unrelated directly to empathy included occasional deviations from correct responses and the models’ short-term memory constraints.

LLMs have shown impressive abilities in semantic understanding and logical reasoning (3). This review supports the idea that LLMs may also demonstrate some abilities that resemble social intelligence. Theory of mind involves the understanding of others thoughts and emotions, and predicting or explaining their behaviors based on these inferences. This concept is fundamental to social interactions, and it is a complex task, as it involves understanding not just the literal meaning of words in a conversation, but the underlying intentions, beliefs, and emotions (18). Several studies evaluated LLMs on theory of mind tasks, with varied performance, depending on the tasks and the models used (18-22).

The definition of empathy varies among researchers and practitioners in social sciences (1). One of the debates is whether it is a cognitive or affective concept, and most definitions of empathy include both (1). *Cognitive empathy* involves the ability to understand another’s feelings, closely related to theory of mind (23). *Affective empathy* relates to experiencing emotions in response to an emotional stimulus (1). The ability of LLMs to demonstrate empathy in various fields as highlighted in this review, seems to align more with the cognitive aspect. It is nevertheless surprising that in some cases the LLM outperformed humans in empathy related tasks.

Research suggests that cognitive and affective empathy are distinct. For instance, people with autism often struggle with cognitive empathy but have normal levels of affective empathy, while psychopathic individuals typically show the reverse pattern (23). Neurological studies demonstrated distinct brain regions associated with each type of empathy, which further supports this notion (24, 25). This differentiation raises questions about the potential evolution of artificial general intelligence (AGI). It is worth questioning if demonstrating cognitive empathy alone is sufficient, or whether affective empathy is imperative for achieving human-like emotional intelligence. If humans cannot distinguish between responses generated by humans and LLMs, or if they prefer AI-generated responses as demonstrated in the study by Ayers et al., perhaps emulating such empathy may be enough.

Numerous studies support the remarkable performance of LLMs in clinical reasoning (4, 5), These models can be applied to enhance the medical care patients receive, while decreasing the workload of healthcare providers (26). Yet, empathy is a key factor in patient care. Empathy in healthcare communication is linked to improved patient satisfaction, adherence to treatment plans, and better outcomes (27). It allows for a more nuanced understanding of patients’ emotional states and experiences, facilitating more compassionate and person-centered care. As such, the ability of LLMs to integrate empathy can significantly enhance the role of AI in healthcare, for both patients and healthcare providers.

This review has several limitations. First, as all but one study evaluated empathy based on subjective assessment, we could not perform a meta-analysis. Second, we only assessed studies directly discussing empathy, while there are many more that evaluate theory of mind tasks that are closely related to “cognitive” empathy. Third, all studies assessed ChatGPT-3.5, and only one study evaluated a model based on LLaMA and GPT-4. This can potentially limit the generalizability of findings to other LLMs. It is possible that alternative LLMs may present different empathy characteristics. Moreover, LLMs are evolving fast, and possibly newer LLMs will present higher cognitive like abilities.

To conclude, this review demonstrates that LLMs exhibit elements of cognitive empathy, being able to recognize emotions and provide emotionally supportive responses in various contexts. Given that social skills are foundational to the concept of “intelligence”, further research is warranted to further develop that aspect in AI. Ultimately, as we continue to refine these models, we approach closer to bridging the gap between artificial and human-like interactions, opening opportunities for empathetic AI applications.

## Data Availability

All data produced in the present work are contained in the manuscript

